## Supplementary material for "Variant-driven multi-wave pattern of COVID-19 via a Machine Learning analysis of spike protein mutations": Methods and supplementary material

[1]Politecnico di Torino, Corso Duca degli Abruzzi 24, 10129 Torino, Italy
[2]Institut de Physique des 2 Infinis (IP2I), CNRS/IN2P3, UMR5822, 69622 Villeurbanne, France
[3]Université de Lyon, Université Claude Bernard Lyon 1, 69001 Lyon, France
[4]Department of Agricultural Sciences, Università degli Studi di Napoli Federico II, Portici, 80055 Italy
[5]Department of Research Infrastructures for Marine Biological Resources (RIMAR), Stazione Zoologica "Anton Dohrn", 80121 Napoli, Italy
[6]Centre International de Recherche en Infectiologie (CIRI), Univ Lyon, Inserm, U1111, Université Claude Bernard Lyon 1, CNRS, UMR5308, ENS de Lyon, 46 Allée d'Italie, 69007 Lyon, France
[7]INFN sezione di Napoli, Complesso Universitario di Monte S. Angelo Edificio 6, via Cintia, 80126 Napoli, Italy
[8]Dipartimento di Ingegneria Università degli studi di Napoli Parthenope, Centro Direzionale di Napoli, Isola C 4, lato Sud, 80143 Napoli, Italy
[9]University of Science and Technology of China (USTC), No.96, JinZhai Road, Baohe District, Hefei, Anhui, 230026, China
[10]Dipartimento di Fisica E. Pancini, Università di Napoli Federico II, Complesso Universitario di Monte S. Angelo Edificio 6, via Cintia, 80126 Napoli, Italy
[11]Scuola Superiore Meridionale, Largo S. Marcellino 10, 80138 Napoli, Italy
[12]CP3-Origins & the Danish Institute for Advanced Study, University of Southern Denmark, Campusvej 55, DK-5230 Odense, Denmark
[*]
[+]these authors contributed equally to this work

#### ABSTRACT

Never before such a vast amount of data, including genome sequencing, has been collected for any viral pandemic than for the current case of COVID-19. This offers the possibility to trace the virus evolution and to assess the role mutations play in its spread within the population, in real time. To this end, we focused on the Spike protein for its central role in mediating viral outbreak and replication in host cells. Employing the Levenshtein distance on the Spike protein sequences, we designed a machine learning algorithm yielding a temporal clustering of the available dataset. From this, we were able to identify and define emerging persistent variants that are in agreement with known evidences. Our novel algorithm allowed us to define persistent variants as chains that remain stable over time and to highlight emerging variants of epidemiological interest as branching events that occur over time. Hence, we determined the relationship and temporal connection between variants of interest and the ensuing passage to dominance of the current variants of concern. Remarkably, the analysis and the relevant tools introduced in our work serve as an early warning for the emergence of new persistent variants once the associated cluster reaches 1% of the time-binned sequence data. We validated our approach and its effectiveness on the onset of the Alpha variant of concern. We further predict that the recently identified lineage AY.4.2 ('Delta plus') is causing a new emerging variant. Comparing our findings with the epidemiological data we demonstrated that each new wave is dominated by a new emerging variant, thus confirming the hypothesis of the existence of a strong correlation between the birth of variants and the pandemic multi-wave temporal pattern. The above allows us to introduce the epidemiology of variants that we described via the Mutation epidemiological Renormalisation Group (MeRG) framework.

### Supplementary material

**S4 Early warning for the AY.4.2 ('Delta plus') driven variant** 10

**S5 3D spike protein analysis** 10

**S6 Heatmap of spike protein variability** 11

**References** 11

### S1 Theoretical modelling of variant diffusion (MeRG)

The current paper deals with the time evolution and spread of different variants of SARS-CoV-2 in a given population. A theoretical study of the underlying processes within the framework of the epidemic Renormalisation Group (eRG) approach[1] has recently been presented in a companion paper[2]. In this section, we briefly review the relevant formalism.

The eRG approach is inspired by the running of fundamental couplings as a function of the energy scale in particle physics. Originally proposed[1] as an effective description of epidemic diffusion processes organised around time-scale invariances, it has been extended to account for geographic mobility across different countries[3], the multi-wave structure of the SARS-CoV-2 pandemic[4] as well as the impact of the US vaccination campaign[5]. The predictive power of this approach has been demonstrated by accurately describing the impact of non-pharmaceutical interventions[4,6] and predicting the starting date of the second wave in the fall of 2020 in Europe[7]. An interpretation of the eRG approach as a time-dependent SIR model has also been discussed[8], while the relation to other epidemiological approaches has been discussed at great depth in this review[9].

In the companion paper[2], the eRG framework has been further extended to describe the time evolution of two different variants of a disease. An epidemic coupling strength $\alpha_i(I_{c,i})$ is introduced for each variant (here $i = 1, \ldots, n$ labels the $n$ different variants). The latter is a function of the cumulative number of individuals $I_{c,i}$ that have been infected by this $i$th variant. The time evolution of the different variants is then described by a set of $\beta$-functions

$$-\beta_i(I_{c,i}) = \frac{d\alpha_i}{dt}\,, \qquad \forall i = 1, \ldots, n\,, \tag{E1}$$

which constitute the core of the Mutation eRG (MeRG) approach. Inspired by the numerical study of a compartmental model and empirically validated by comparing with data from California, the United Kingdom and South Africa, the $\beta$-functions were written in the form of gradient equations[2]

$$-\beta_i(I_{c,i}) = \nabla_i \Phi(I_{c,j})\,, \qquad \text{with} \qquad \begin{array}{l} \nabla_i = \frac{\partial}{\partial I_{c,i}} \\ \Phi(I_{c,j}) = \sum_{k=1}^n I_{c,k}^2 \frac{\gamma_k}{2}\left(1 - \frac{2 I_{c,k}}{3 A_k}\right)\,. \end{array} \tag{E2}$$

Here, $\gamma_k$ is a measure for the infection rate and $A_k$ is the asymptotic number of individuals infected by variant $k$. Solutions of the $\beta$-functions (E2) give cumulative numbers of infected individuals as functions of time for each variant that follow a logistics function

$$I_{c,i}(t) = \frac{A_i}{1 + e^{-\gamma_i(t-\kappa_i)}}\,, \tag{E3}$$

where $\kappa_i$ is a parameter that governs the time of appearance of the variant. For given $i$, the function $\beta_i$ in (E2) has two zeroes, namely $I_{c,i} = 0$ and $I_{c,i} = A_i$, corresponding to the complete absence of the variant $i$ or the eradication of the latter, in the sense that there are no more infectious individuals left carrying it. The complete set of $\beta$-functions $(\beta_1, \ldots, \beta_n)$ has $2^n$ fixed points and the epidemic is described by the flow equations connecting (some of) them.

#### S2 Machine Learning algorithm

We employ a machine learning algorithm based on the Levenshtein distance between spike protein sequences. The procedure can be grossly divided into three steps, which we describe in detail in the next three sub-sections S2.1–S2.3. All the python codes are provided (see link in the main publication), with reference to the main libraries provided in this material. While in this work we mainly focus on England, due to the larger dataset, and other nations of Great Britain, the ML codes can be run on any dataset, for different countries or regions of the world.

The spike protein sequencing data for the UK nations were collected from the GISAID repository[10,11]. For each nation we analyse the data via a simple Machine Learning (ML) algorithm based on the Levenshtein measure (LM)[12,13], which quantifies the difference between two strings of characters. This ML approach has been long used in biology, with appropriate refinements and optimisations[14], while more recently deep learning approaches[15–17] are becoming more effective.

##### S2.1 Extraction and pruning of the raw data

The raw data are downloaded from the GISAID open-source repository (registration required) in the form of "fasta" files, which contain information on samples from COVID-19 infected cases. The files contain the full genome, including the spike protein sequences, but also the date when the sample was taken, the laboratory where it was analysed and the geographic information about the country or region of origin of the sample. This additional information allows to separate datasets based on a specific geographical origin, and sample them in time.

In this work, we focus only on the spike protein sequences. The data contains sequences with un-identified amino-acids (labelled with an X) and sequences with missing pieces (thus, with unusual lengths). The spike protein of the early SARS-CoV-2 sequences have 1271 amino-acids. Hence, to remove data with missing pieces, we only keep sequences with at least 1250 amino-acids. Furthermore, we dismiss all sequences containing at least one X in the sequence. This pruning allows us to work only on a high purity dataset. The number of sequences before and after the pruning for England, Wales and Scotland are listed in Table T1. After the pruning, a significant number of sequences are in the dataset, many of which contain the same sequences for the spike protein. To accelerate the next step in the analysis, it is convenient to remove repetitions and thus only work on a dataset containing only different sequences (see right column in Table T1). In particular, this is necessary when we do a global analysis of the whole dataset, while the time-sampling automatically reduces the number of sequences in the dataset.

| | Raw sequences | After pruning | Different sequences (after pruning) |
|---|---|---|---|
| England | 646.697 | 461.122 | 13.887 |
| Wales | 44.944 | 26.440 | 1.315 |
| Scotland | 64.409 | 47.137 | 1.936 |

**Table T1.** Number of sequences in the datasets for England, Wales and Scotland before and after the pruning and selection.

The extraction and pruning of the data is done by the python program `extraction_country.py`, where the name of the country or region needs to be specified in the first lines of the program. The output is as follows:

- The list of the strictly different sequences as a text file `country_seq_ass.txt`.
- A csv table `country.csv` were lines correspond to selected sequences. The first column contains the date, the second the corresponding sequence in reference to the text file already saved and finally a column labelling the VoC or VoI the sequence belongs to, according to GISAID.
- A list of the different labs contributing to the sequencing.

Note, in passing, that the pruning could be by-passed by modifying the distance calculation in such a way that the presence of incomplete sequences is properly taken into account. This would require a more complex and optimised procedure. For our purposes, we wanted to remain as unbiased as possible, thus we opted for the pruning and decided to work with a dataset of the highest purity.

##### S2.2 Computation of the distance matrix

The output of the previous step is a list of strictly different spike protein sequences, with a record of their multiplicity in the dataset under study. At this stage, we need to compute the Levenshtein distances between them. This computation yields a symmetric matrix with zeros on the diagonal, such that only a triangular matrix needs to be computed on the data.

To efficiently compute the Levenshtein distances, we use the library polyleven. For this purpose, we created the python code `Launch_distance.py`, where a specific country or region has to be specified at the beginning of the file. The program calls

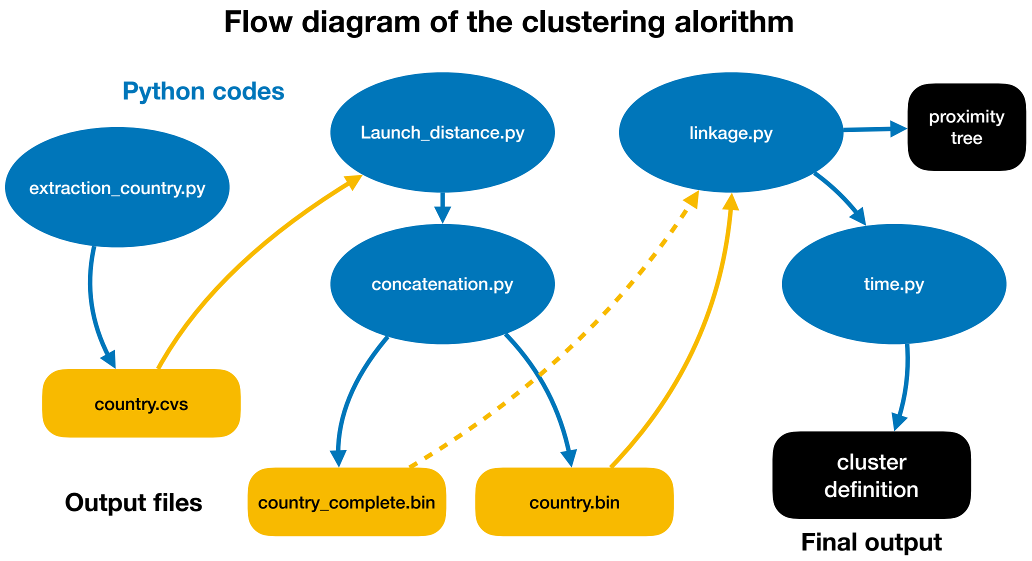

**Figure F1.** Flow diagram of the clustering algorithm, indicating the concatenation of python programs (in blue) and intermediate output/input files (in yellow). The string "country" needs to be replaced by the name of the country under study, for instance "England".

`distance.sh` and `distance.py`, thus the process is multicore and very fast. The various lines of the upper triangular matrix are saved in the subfolder `create` as separate binary files. Launching now `concatenation.py` will load them all and save a single file containing the complete distance matrix, while the temporary files are deleted.

Note that to speed up the computation of the distances, we only considered strictly different spike protein sequences. This avoids repeated computations. Yet, the construction of the proximity tree, which will be the task of the next step, may depend on the multiplicity of each sequence in the starting dataset, as we will see. Hence, the program `concatenation.py` outputs two different distance matrices: `country.bin` where only strictly different sequences are included, and `country_complete.bin` where the multiplicity of the sequences are taken into account by copying multiple times the corresponding lines in the table. Thus, the latter contains a much bigger distance matrix than the former.

##### S2.3 The proximity tree

To create the proximity tree, various libraries are at our disposal. We chose to adopt the hierarchical clustering algorithm from the `scipy` library as it is easy to work with and adaptable. The only needed input is the distance matrix. For the algorithm, each initial sequence, i.e. a line or column of the distance matrix, is a *leaf*. The library then groups the leaves into *branches* based on the Levenshtein distance between them, as specified in the input file. To calculate the effective distance between branches, various methods are available, and they need to be selected. We anticipate that some methods are not sensitive to the multiplicity the identical leaves (which have distance zero among them), while others are.

The algorithm constructs the proximity tree starting from the leaves: hence, for a sample with $n$ leaves (i.e. a $n \times n$ distance matrix as input), the code considers an initial state with $n$ branches containing a single leaf each. Hence, the steps consist in regrouping 2 branches together forming a new branch. Naturally, there can be at most $n-1$ of those steps. At any step, the two branches that are chosen to be regrouped (let us call them $A$ and $B$) are the ones that have the smallest distance between them. Such distance $\mathrm{dis}(A,B)$ is computed in terms of the Levenshtein distance $d(x,y)$ between leaves on the two branches ($x$ and $y$ being a leaf from $A$ and $B$ respectively). Here is where different methods to compute the branch distance $\mathrm{dis}(A,B)$ can be employed. The most common choices are:

- Single Linkage Clustering, where

$$\mathrm{dis}(A,B) = \min_{x\in A\,,\, y\in B} \;\; d(x,y)\,. \tag{E4}$$

  This method is insensitive to the multiplicity of identical leaves, thus one can use the file `country.bin` as input.

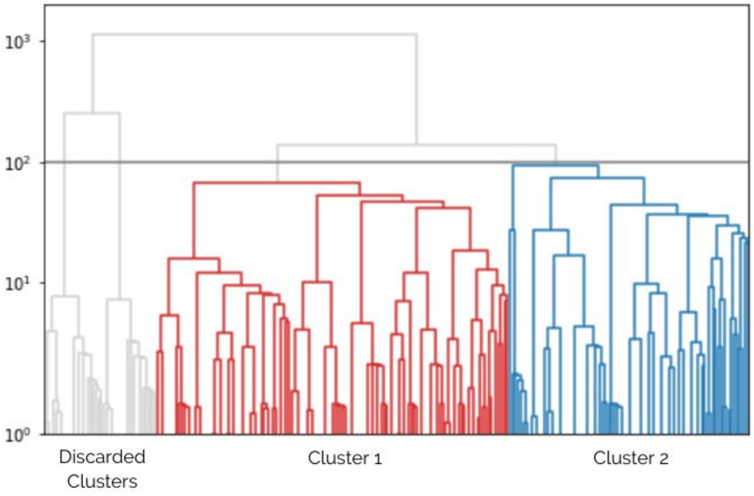

**Figure F2.** Dendrogram three for the England dataset, for month 9 (September 2020), built using the Ward's method. On the y-axis we show the effective distance between two clusters at the point when they merge. The branches in colour represent more than 1% of the total number of sequences for a cut at Ward distance 100.

- Complete Linkage Clustering, where

$$\mathrm{dis}(A,B) = \max_{x \in A\,,\, y \in B} \; d(x,y)\,. \tag{E5}$$

As before, this is independent on the leaf multiplicity.

- Unweighted Average Linkage Clustering, where

$$\mathrm{dis}(A,B) = \frac{1}{|A||B|} \sum_{x \in A\,,\, y \in B} d(x,y)\,, \tag{E6}$$

where $|X|$ is the number of leaves in the branch $X$. This method is sensitive to the multiplicity of identical leaves, thus is requires `country_complete.bin` as input.

- Ward's Method, an agglomerative clustering method based on the measure of the average squared distance of points in the cluster to its centre of gravity, or centroid. Hence, the effective distance between two branches is defined by the increase in the above measure in the merged cluster with respect to the two separate ones. In practice:

$$\mathrm{dis}(A,B) = \frac{|A||B|}{|A|+|B|} \left[ \sum_{x \in A\,,\, y \in B} \frac{d(x,y)^2}{|A||B|} - \sum_{x,x' \in A} \frac{d(x,x')^2}{2|A|^2} - \sum_{y,y' \in B} \frac{d(y,y')^2}{2|B|^2} \right]. \tag{E7}$$

This method is sensitive to the multiplicity of identical leaves, thus it requires `country_complete.bin` as input. Note that we also used this method on the distance matrix `country.bin` to speed up the computation.

At each step, the algorithm joins branches together, until all leaves are grouped together. This approach can be schematically represented by a dendrogram tree, as shown in Fig. F2. This step is executed by the program `linkage.py`. In order to define clusters and variants, we need to define a threshold in the effective distance, which will therefore be equivalent to a horizontal cut of the tree branches. In practice, all clusters whose effective distance is larger than the threshold are used to define the variants. The value of the threshold needs to be determined each time, as it crucially depends on the measure that is employed, and on the samples. Once the threshold is fixed, the clusters and their time evolution can be obtained. This is done by the executable called `time.py`.

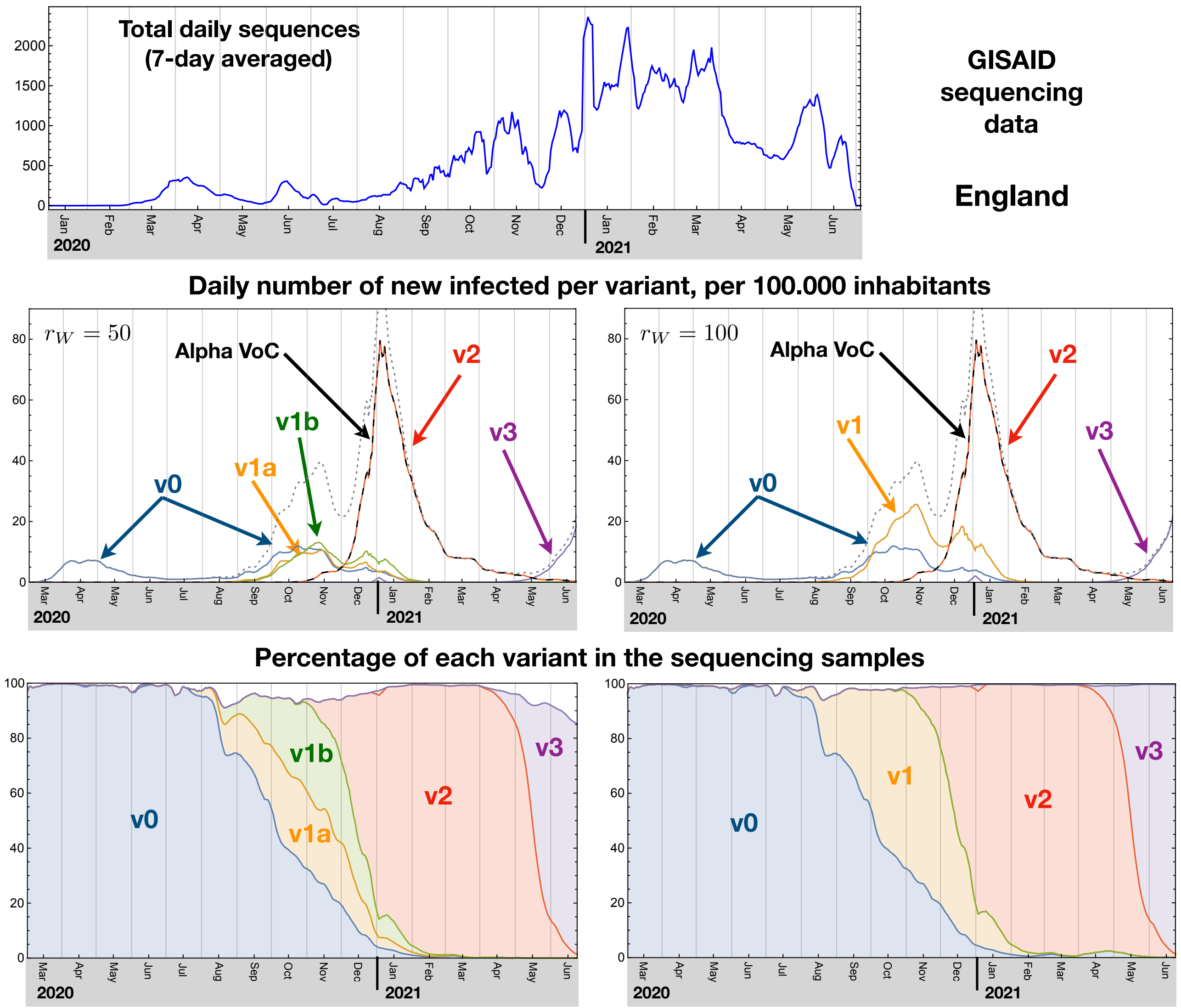

**Figure F3.** **Machine Learning results.** For England, the top panel shows the time-distribution of the sequences used in this analysis. Below, we present the results of the clustering for two choices of cut: at a Ward distance $r_W = 50$ (left) and $r_W = 100$ (right). The middle plots show an estimate of the daily number of new infections per cluster, compared to the ones attributed to the Alpha VoC in the GISAID dataset dotted and solid lines, respectively. In the bottom plots, we show the evolution of the percentage of sequences in each cluster.

##### S2.4 Application to the England data set

As a first step, we apply the ML algorithm to all sequences in the England dataset, updated to June 2021. The clustering is done on the data that contains only different sequences for the spike proteins to reduce the number of data points, as contained in the file `england.bin`. After defining the clustering, the number of total sequences in each cluster is counted and each sequence time-tagged, in order to obtain the frequency for the occurrence of each cluster as a function of time. The results are shown in Fig. F3 for two choices of the cut in the Ward distance: $r_W = 50$ in the left plots and $r_W = 100$ in the right plots. We consider as relevant clusters only the ones that contain at least 1% of the sequences within the full dataset. We show two choices of $r_W$ within the defined working region to illustrate how lowering or increasing the threshold leads to splitting or merging clusters. We identify, therefore, 5 clusters for the lower cut (v0, v1a, v1b, v2, v3), and 4 for the upper cut (v0, v1, v2, v3). Increasing the cut allows to merge the clusters v1a and v1b into v1. We also checked that by increasing the $r_W$ cut, it is v0 and v1 ($r_W = 100$) that merge into a single cluster, while v3 remains separated. This analysis can give a rough idea of the closeness of different

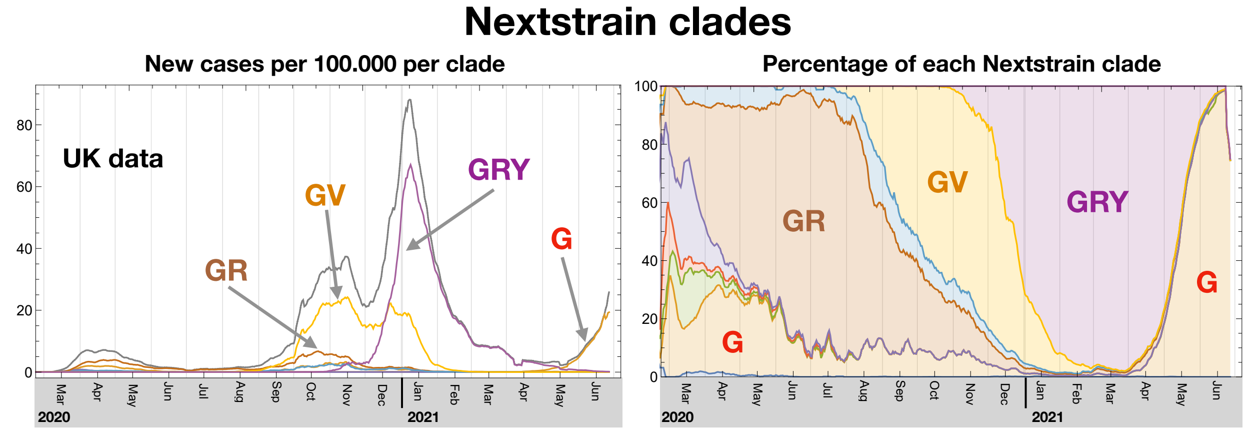

**Figure F4. Nextstrain clades.** Representation of the "clades" as defined by the Nextstrain initiative[18], where we only label the most frequent ones, G, GR, GV and GRY. In the two panels we show the estimate of the number of infected by each clade (left) and the percentage in the sequencing data (right) for the UK. We indicate the names of only the major clades. Note that GRY coincides with the Alpha VoC, while the Delta VoC is behind the late dominance of the clade G.

clusters via the merging/splitting due to the change in the clustering threshold. A more detailed analysis of the phylogeny of the variants, and their connection, is presented in the main text via a time-ordered clustering, as described in detail in the next section.

In the middle and bottom plots of Fig. F3 we report our estimate of the number of daily new infections in each clustered variant, computed by multiplying the measured frequency in the sequencing dataset with the reported number of new infections, and the frequency of appearance of each cluster in the analysed dataset. These results can be compared to the ones in Fig.2 of the main publication, even though the method to obtain them is different, as well as the values of the thresholds used to define the clusters. This comparison validates the solidity of the time-ordered analysis showed in this article. The results also corroborate the idea that each wave can be associated with a different dominant cluster (as defined at $r_W = 100$): v0 for the first wave occurring in March-May 2020; v1 for the second wave in October-November 2020; v2 (the Alpha VoC) for the third wave in December-February 2021; and v3 for the last wave starting in May-June 2021.

Before further analysing the data, we validated our method by comparing the variant definition, associated to the clusters in our analysis, to more standard methods used in computational biology. For this purpose, we have chosen the clade classification[19], as defined by the Nextstrain initiative[18] and embedded in the data from the GISAID repository. The Nextstrain clade definitions are informed by the statistical distribution of genome distances in phylogenetic clusters[20], followed by the merging of smaller lineages into major clades. The latter is based on shared marker variants. The main difference compared to our analysis is that the comparison is based on the whole genome sequence, while we only analyse spike proteins. Note that the marker variants used in the lineage merging contain specific information on proteins, including the spike. It has been noted that this way of defining clades provides similar results to the Pango lineage classification[21], and other variations. For the UK, we show in Fig. F4 the frequency and number of infections assigned to each clade, where we note that GRY corresponds broadly to the Alpha VoC (or variant v2 in our results). These plots confirm that each wave is dominated by a single group of mutations. By comparing the frequencies, we also observe that our variant v1 matches the clade GV, while v0 groups all the other ones that mainly occurred during the first wave. We also note that our method allows to clearly identify the Delta VoC (v3), while in the Nextstrain clades it is associated with the clade G. This validation demonstrates that the clustering based on the spike protein sequences alone is able to identify relevant variants for the SARS-CoV-2.

#### S3 Time-binning and linkage analysis

Data from the GISAID repository has been binned on a monthly basis for the England, Wales and Scotland datasets, as shown in Table T2. Despite the rather large number of monthly recorded data, we found sequences with high replica rate, thus the percentage of different sequences over the whole dataset is also reported. For each month the same hierarchical clustering algorithm described above based on the Levenshtein measure (LM) has been applied. Here the two main parameters to take in account are the clustering cutoff that mainly acts on the size of the clusters and the threshold on the minimum amount of data that is needed to define a cluster. Detailed studies have been made to find the right trade off between the number of clusters and the coverage of the dataset.

| Month | England | Scotland | Wales |
|---|---|---|---|
| 1 | 2 (100%) | 0 | 0 |
| 2 | 64 (9%) | 0 | 1 (100%) |
| 3 | 3944 (5%) | 1109 (5%) | 965 (5%) |
| 4 | 7335 (6%) | 1923 (6%) | 2178 (5%) |
| 5 | 2204 (10%) | 306 (12%) | 866 (7%) |
| 6 | 5079 (6%) | 58 (24%) | 425 (7%) |
| 7 | 2026 (9%) | 67 (9%) | 102 (11%) |
| 8 | 4429 (8%) | 937 (8%) | 210 (15%) |
| 9 | 9499 (6%) | 1592 (6%) | 1275 (6%) |
| 10 | 19634 (6%) | 1740 (8%) | 2732 (5%) |
| 11 | 23431 (7%) | 602 (11%) | 2497 (6%) |
| 12 | 25339 (6%) | 1161 (9%) | 3989 (6%) |
| 13 | 46685 (5%) | 2196 (8%) | 3508 (6%) |
| 14 | 39326 (5%) | 4600 (6%) | 2769 (7%) |
| 15 | 49545 (4%) | 9655 (3%) | 1958 (7%) |
| 16 | 24057 (5%) | 4800 (5%) | 689 (12%) |
| 17 | 24335 (4%) | 6263 (3%) | 301 (10%) |
| 18 | 30981 (4%) | 3638 (5%) | 98 (12%) |
| 19 | 58600 (4%) | 4968 (9%) | 1580 (12%) |
| 20 | 86520 (5%) | 1522 (15%) | 297 (25%) |

**Table T2.** Number of GISAID recorded sequences from January 2020 (Month 1) to August 2021 (Month 20).

We define the cutoff based on the Ward distance, $r_W$, as defined in the previous section. The coverage threshold is defined in terms of a minimum percentage of the whole sequence dataset (per month) that is covered by each branch above the cutoff. Only branches above the threshold are retained to define clusters. For each given cutoff value $r_W$, the coverage of the dataset decreases while the threshold value increases. Hence, increasing the coverage would require pushing the threshold towards smaller values. However, the threshold choice must take into account the large increase in the number of (small) clusters. Quite independently of the month, the number of clusters is almost stable for any coverage threshold in the $[10^{-2}, 10^{-1}]$ range, while for smaller values it shows a linear increase. Thus we choose to set the threshold value to $10^{-2}$ (1%) to maximise the mean coverage of the dataset (mean coverage $> 90\%$). Moreover it has been found that the mean coverage is almost stable for any cutoff in the range $r_W \in [50, 200]$ while the number of defined clusters decreases with the cutoff value, as shown in Fig. F5. Thus the default working point has been set to a cutoff value of $r_W = 100$ and a coverage threshold of 1%.

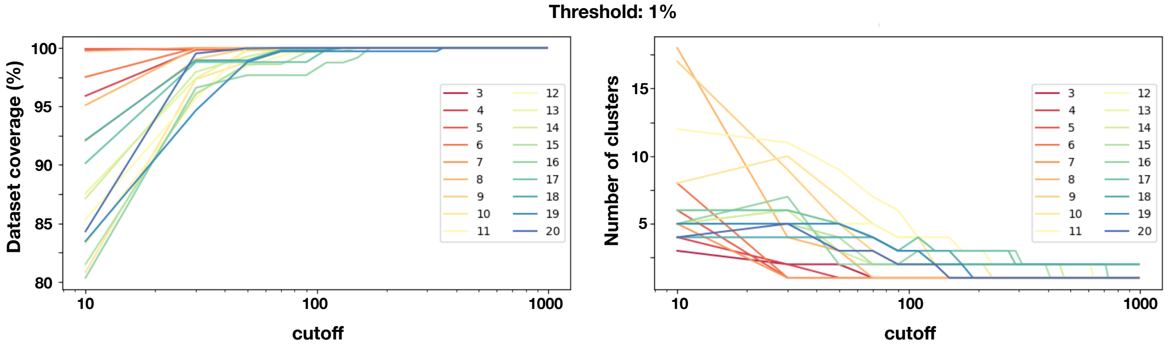

**Figure F5.** Dataset coverage (left) and number of clusters (right) as a function of the Ward cutoff $r_W$.

##### S3.1 Time-series sequences

To define and follow the time evolution of a given candidate variant, we build time-series of clusters starting from month 1 using the following algorithm:

- For each cluster in month $i$ we selected the most frequent sequence and we tried to find a cluster with the same sequence in month $i+1$ (**strong link**). We iterate to build a path until the procedure fails or the last month is reached.

- If the strong link association fails but we still have clusters in consecutive months that are free from strong links, we connected them provided that the distance is less than a given threshold. If two clusters converge to the same node we keep the nearest one (**weak link**).

The threshold used to define weak links has been optimised looking at the distribution of the distances between clusters connected with strong links, as shown in Fig. F6. To preserve the topology based on the LM, we chose the soft link threshold as the maximum distance found for a strong link.

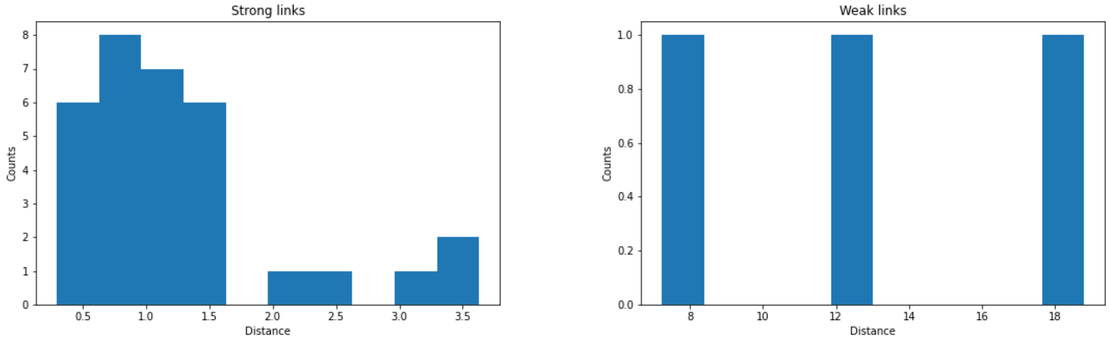

**Figure F6.** Strong link (left) and weak link (right) distances.

We define a chain as a list of consecutive clusters connected by strong or weak links as best candidate to study the evolutionary paths of a candidate variant. For each well defined chain we also assign a branching link taking the first cluster of the chain and associating it with the cluster in the previous month that is the closest in terms of the Ward distance. In the following we assign as chain identification number the one of the first cluster in the chain.

##### S3.2 Results for the nations of Great Britain: England, Scotland and Wales

Chains of clusters for England have been presented in the main manuscript, at the top of Fig.2. We also repeated the same analysis for Scotland and Wales datasets, even though the statistics is more limited. Using the same working point, with cutoff $r_W = 100$ and threshold of 1%, comparable with the right column of Fig.2, we obtain the results shown in Fig. F7. For both nations, we can only define three persisting chains: The chains 1–15 (Scotland) and 1–19 (Wales) can be associated to the original variant of the virus (blue solid lines); the chains 11–21 (Scotland) and 13–24 (Wales) contain as dominant Spike variant the one with the characteristics of the Alpha VoC, B.1.1.7 (yellow solid lines); the chains 18–23 (Scotland) and 22-25 (Wales) share the dominant Spike variant of the Delta VoC, B.1.617 with the additional, mutation T95I (solid green lines). These results validate the ability of our ML algorithm to identify emerging variants.

##### S3.3 Virus diversification along the chains for England

A comparison of the evolutionary path of the England chains suggests a competitive mechanism between variants. To further investigate this, we studied the stability of the chains via the distance between consecutive clusters, as shown in Fig. F8. For longer chains, like v0 (original variant) and v2 (Alpha VoC), we clearly see that the distance increases towards the end of the chain itself. Furthermore, v2 enjoys smaller distances along the chain, thus showing increased stability.

For completeness, we also provide some detailed information about the chains, also listing the frequencies of the dominant and sub-dominant Spike variants in all the chains for England, as shown in Table T4.

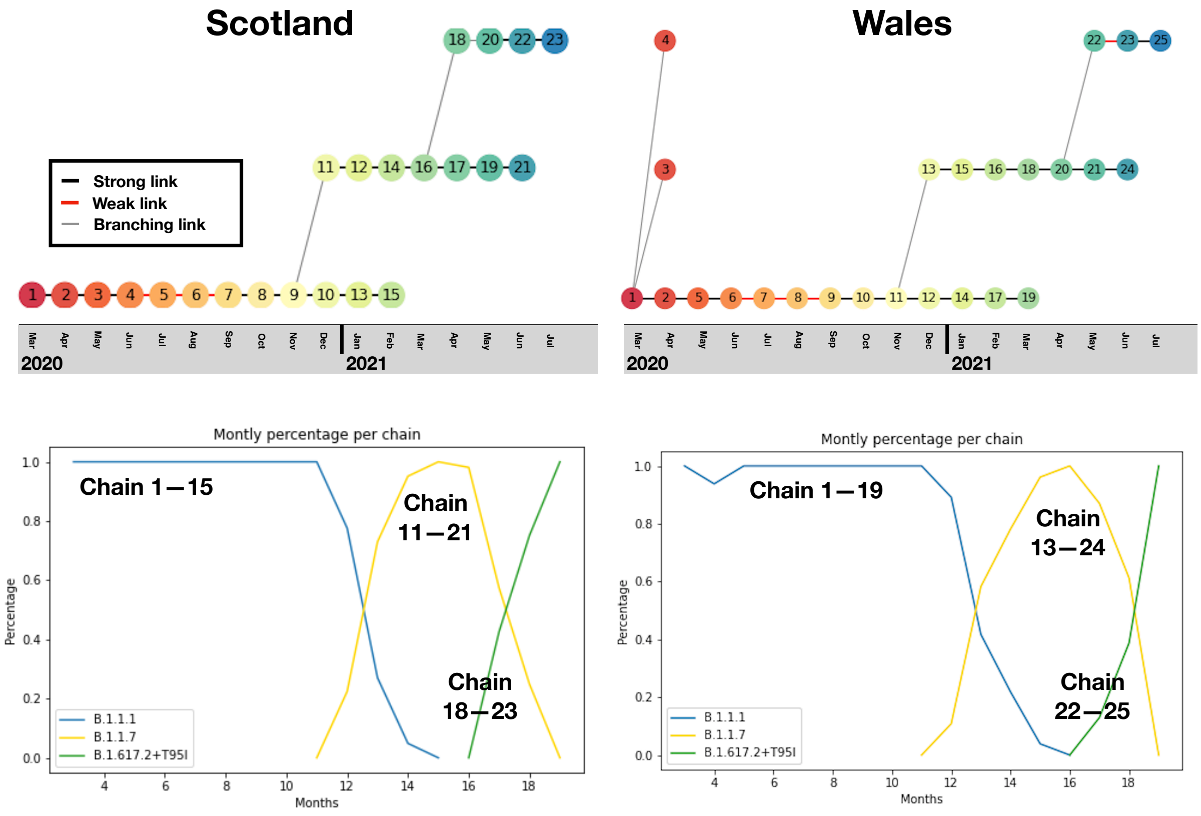

**Figure F7.** Monthly chains as candidate emerging variants for the Scotland dataset (left) and the Wales one (right). Strong links (black line) and weak link (red line) are shown within each chain. In the bottom plots we show the percentage of each chain per month.

#### S4 Early warning for the AY.4.2 ('Delta plus') driven variant

We performed a preliminary weekly-binned analysis for the most recent data (July to September, 2021) to highlight the emergence of a new variant. In particular, we aim at uncover if the branching observed in Fig.2 of the main text in July/August can trigger an early warning issue. The data is summarised in Table T3, where we highlight the sequences belonging to the Pango lineage AY.4.2, which is the dominant Spike variant in cluster 43 of Fig.2. In this preliminary study, we applied the ML algorithm with the working point $r_W = 100$ and threshold of 1%, before the optimisation for the weekly analysis.

The resulting chains are shown in Fig. F9, where the cluster 20 and the chain 25–28–30 have the AY.4.2 as dominant Spike variant. The lack of a cluster in week 36 may be due to the lower statistics, and it can be filled via the optimisation of the working point. These preliminary results clearly show that a new persisting chain is emerging, starting from week 35 (23rd of August, 2021). Note that our identification predates the establishment of the lineage itself, which was done at the beginning of September 2021.

#### S5 3D spike protein analysis

3D structures of the closed Spike conformation of SARS-CoV2[22] (PDB 6ZGI) were analysed using the UCSF ChimeraX software, developed by the Resource for Biocomputing, Visualisation, and Informatics at the University of California, San Francisco, with support from National Institutes of Health R01-GM129325 and the Office of Cyber Infrastructure and Computational Biology, National Institute of Allergy and Infectious Diseases[23].

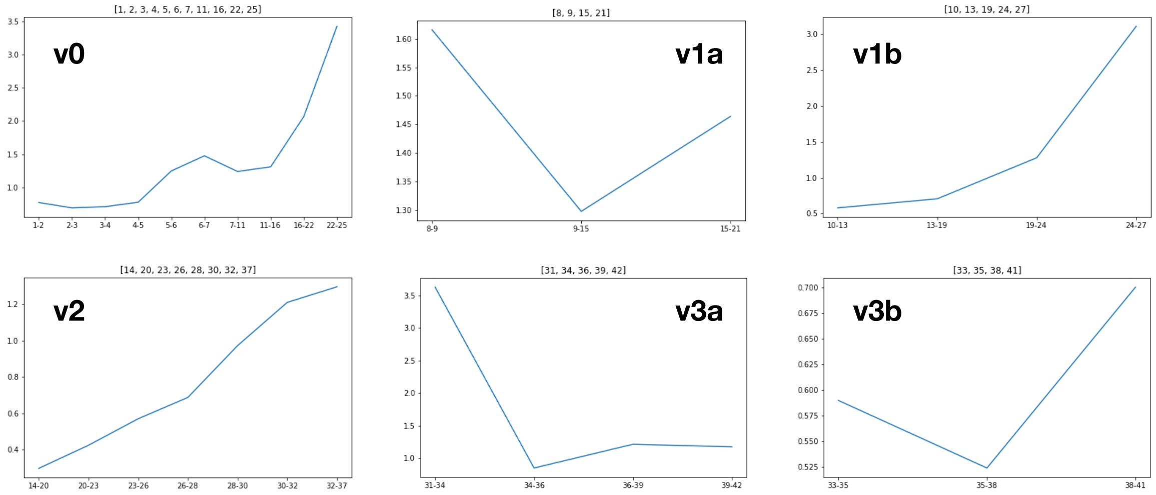

**Figure F8.** Ward distance between consecutive clusters for the England chains ($r_W = 100$): v0 (original strain, upper left), v1a (upper middle), v1b (upper right), v2 (Alpha VoC, lower left), v3a (Delta VoC, lower middle) and v3b (Delta VoC, lower right)

#### S6 Heatmap of spike protein variability

For each time-ordered chain, it was possible to investigate the local path of mutations along time. As the number of sequences in each cluster varies a lot, depending on the month they refer to due to variability in the data collection, we need to find a way to properly normalise the counting of mutations at each position. To do this we applied the following procedure: We first define a vector counter (dimension = 1273, the number of the sites in the spike protein) and initialised it to 0. Our goal is to increase this counter in the sites in which a mutation occur. To do so, we apply the following algorithm:

1. For each cluster, we select the sequences repeated more than 1% of the total elements in the cluster.
2. For each sequence in the cluster $i$, we determine the sequence in cluster $i-1$ of the chain that is at the minimal Levenshtein distance. This procedure defines a pair of sequences, Seq1 in cluster $i$ and Seq2 in cluster $i-1$.
3. We compare the two sequences and, if we find a mismatch of amino acid in the same position, we increment the counter in that position taking into account the number of sequence Seq1.
4. This procedure is repeated for all the sequences selected in cluster $i$
5. At the end, once all sequences in cluster $i$ are exhausted, we divide the counter by the number of all elements present in the cluster.

For the first cluster in each chain, we used as reference the dominant sequence (variant) in the the paren cluster, as defined by the branching links. For v0, we used as reference the sequence from Wuhan to compute the mismatch for the cluster 1.

| Week (2021) | Tot. sequences | AY.4.2 sequences | percentage |
|---|---|---|---|
| 26 | 10942 | 28 | 0.25% |
| 27 | 11133 | 78 | 0.7% |
| 28 | 10202 | 121 | 1.2% |
| 29 | 16120 | 221 | 1.4% |
| 30 | 17234 | 248 | 1.4% |
| 31 | 19654 | 309 | 1.6% |
| 32 | 16665 | 380 | 2.3% |
| 33 | 19886 | 547 | 2.75% |
| 34 | 20051 | 683 | 3.4% |
| 35 | 20262 | 772 | 3.8% |
| 36 | 13503 | 591 | 4.4% |
| 37 | 18009 | 1045 | 5.8% |
| 38 | 15609 | 1074 | 6.9% |
| 39 | 9327 | 757 | 8.2% |

**Table T3.** Number of GISAID recorded sequences from week 26 (20th of June, 2021) to week 39 (19th of September, 2021) indicating the number of sequences and percentage of the Pango lineage AY.4.2.

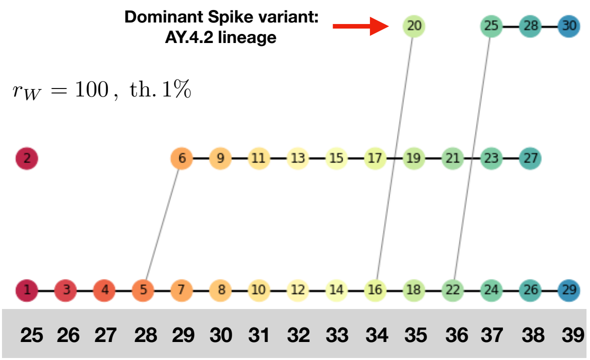

**Figure F9.** Results of the clustering algorithm for the weekly data, highlighting the emergence of a new persisting chain (red arrow) dominated by the AY.4.2 lineage. The working point is $r_W = 100$ and threshold of 1%.

| Month | Cluster | sequences | n$^{o}$ variants | Spike variant percentage |
|---|---|---|---|---|
| Mar 2020 | 1 | 3571 | 3 | (60%, 37%, 2.7%) |
| Apr 2020 | 2 | 6218 | 4 | (84%, 13%, 1.8%, 1.2%) |
| May 2020 | 3 | 1744 | 6 | (89.5%, 3.3%, 2.5%, 2.2%, 1.3%, 1.3%) |
| Jun 2020 | 4 | 3833 | 5 | (89%, 6.1%, 1.9%, 1.7%, 1.5%) |
| Jul 2020 | 5 | 1504 | 8 | (83%, 5.8%, 3.4%, 1.9%, 1.7%, 1.6%, 1.5%, 1.3%) |
| Aug 2020 | 6 | 3288 | 14 | (56.5%, 11%, 5.0%, 4.1%, 3.5%, 3.4%, 3.0%, 2.7%, 2.3%, 2.3%, 1.9%, 1.9%, 1.3%, 1.3%) |
| Sept 2020 | 7 | 4795 | 12 | (69%, 7.4%, 3.6%, 3.3%, 2.8%, 2.5%, 2.4%, 2.3%, 1.9%, 1.9%, 1.6%, 1.3%) |
| | 8 | 3012 | 10 | (42%, 33.6%, 5.6%, 4.9%, 3.7%, 3.2%, 1.9%, 1.9%, 1.7%, 1.5%) |
| Oct 2020 | 9 | 5410 | 9 | (67%, 8.7%, 6.7%, 4.8%, 2.9%, 2.7%, 2.5%, 1.9%, 1.5%) |
| | 10 | 4517 | 6 | (89.5%, 3.6%, 2.1%, 2.0%, 1.4%, 1.3%) |
| | 11 | 5223 | 8 | (75%, 10.5%, 3.0%, 2.8%, 2.6%, 2.4%, 1.9%, 1.7%) |
| | 12 | 389 | 4 | (60.7%, 16.5%, 16%, 6.7%) |
| Nov 2020 | 13 | 6703 | 6 | (86.4%, 4.7%, 3.9%, 2.0%, 1/6%, 1.3%) |
| | 14 | 1762 | 5 | (93$, 3.0%, 1.4%, 1.2%, 1.2%) |
| | 15 | 5080 | 6 | (74.7%, 12%, 4.2%, 4.2%, 2.5%, 2.4%) |
| | 16 | 3699 | 8 | (70.5%, 11%, 6.2%, 3.1%, 3.0%, 2.7%, 2.0%, 1.5%) |
| | 17 | 953 | 10 | (48.4%, 13.4%, 12.7%, 10%, 5.8%, 3.5%, 1.9%, 1.8%, 1.5%, 1.2%) |
| | 18 | 260 | 8 | (68%, 12.7%, 6.5%, 4.2%, 2.7%, 2.7%, 1.9%, 1.2%) |
| Dec 2020 | 19 | 3951 | 5 | (89%, 3.7%, 3.0%, 2.6%, 1.9%) |
| | 20 | 12153 | 2 | (98.6%, 1.4%) |
| | 21 | 2996 | 9 | (63.8%, 9.5%, 6.4%, 5.4%, 5.0%, 3.4%, 2.7%, 1.9%, 1.8%) |
| | 22 | 1204 | 11 | (60%, 10.8%, 9.5%, 3.9%, 2.9%, 2.3%, 2/3%, 2.1%, 2.1%, 2.1%, 2.0) |
| Jan 2021 | 23 | 33238 | 3 | (95.2%, 3.3%, 1.5%) |
| | 24 | 3287 | 7 | (64%, 22.3%, 5.4%, 2.8%, 2.2%, 1.9%, 1.6%) |
| | 25 | 464 | 17 | (33.8%, 8.4%, 7.3%, 6.7%, 6.3%, 5.8%, 4.5%, 4.1%, 4.1%, 3.2%, 2.6%, 2.6%, 2.4%, 2.4%, 1.9%, 1.9%, 1.9%) |
| Feb 2021 | 26 | 29677 | 5 | (92.7%, 2.4%, 1.9%, 1.6%, 1.5%) |
| | 27 | 548 | 9 | (51.6%, 17%, 10.4%, 8.4%, 3.6%, 3.3%, 2.0%, 1.8%, 1.8%) |
| Mar 2021 | 28 | 36439 | 5 | (91.2%, 2.7%, 2.4%, 2.1%, 1.5) |
| | 29 | 358 | 16 | (25.7%, 15.4%, 10.6%, 6.1%, 5.6%, 5.3%, 5.0%, 5.0%, 4.5%, 3.4%, 2.5%, 2.5%, 2.2%, 2.2%, 2.0%, 2.0%) |
| Apr 2021 | 30 | 16020 | 4 | (93.4%, 2.9%, 1.9%, 1.7%) |
| | 31 | 819 | 12 | (34.9%, 21.2%, 17.8%, 8.5%, 5.0%, 3.5%, 2.1%, 1.7%, 1.3%, 1.3%, 1.2%, 1.2%) |
| May 2021 | 32 | 7317 | 5 | (90.5%, 3.0%, 3.0%, 1.9%, 1.6%) |
| | 33 | 8673 | 2 | (92.4%, 7.6%) |
| | 34 | 3972 | 3 | (62%, 35.4%, 2.6%) |
| Jun 2021 | 35 | 18918 | 2 | (98.8%, 1.2%) |
| | 36 | 5690 | 2 | (63.2%, 36.8%) |
| | 37 | 1367 | 5 | (93.3%, 2.0%, 1.8%, 1.5%, 1.4) |
| Jul 2021 | 38 | 35501 | 2 | (98.4%, 1.4%) |
| | 39 | 7627 | 7 | (88.2%, 3.1%, 2.2%, 2.2%, 1.5%, 1.5%, 1.4) |
| | 40 | 2078 | 4 | (68.3%, 29%, 1.3%, 1.3%) |
| Aug 2021 | 41 | 70724 | 2 | (67.8%, 1.2%) |
| | 42 | 13548 | 5 | (66.2%, 5.0%, 3.6%, 1.7%, 1.3%) |
| | 43 | 2248 | 5 | (78.9%, 6.5%, 1.8%, 1.2%, 1.1%) |

**Table T4.** Dominant and subdominant Spike proteins in the England clusters from March 2020 to August 2021.
